# Causal effects of lung function and COPD on functional outcome following ischemic stroke: a univariable and multivariable Mendelian randomization study

**DOI:** 10.1101/2025.09.03.25335062

**Authors:** Wan-ting Sun, Juan Wang, Zhuo-ya Yao, Fang Pan, Zhao-dan Gan, Yun Yang, Jun Lu, Long-yun Zhou, Guang-xu Xu

## Abstract

**Background and Objectives:** Studies have shown that lung function and chronic obstructive pulmonary disease (COPD) are associated with functional outcome for several diseases, as well as the onset of stroke. However, the causal relationship between lung function, COPD and the functional outcome of ischemic stroke (IS) is unclear. The aim of this study was to investigate the causal relationship between lung function, COPD and IS functional outcomes using Mendelian randomization (MR).

**Methods:** Lung function data were derived from the Genome-wide Association Study (GWAS) summary database. Data of COPD were derived from the national genetic factors data repository of the FinnGen project in Finland. The outcome data of IS function were obtained from the Genetics of Ischemic Stroke Functional Outcome (GISCOME) Network. Univariate MR analysis mainly used the inverse variance weighting (IVW) method, MR-Egger, weighted median method and two-sample Maximum Likelihood to evaluate the causal effects of lung function and COPD on the prognosis of IS. Sensitivity analyses were conducted by evaluating heterogeneity and potential pleiotropy. Additionally, multivariable MR analyses were employed to ascertain whether these effects were independent.

**Results:** In univariate MR analyses, the estimates of IVW, MR-Egger, weighted median method and two-sample Maximum Likelihood analysis all showed that there was no genetic causal relationship between COPD, FVC, FEV1, FEV1/FVC, PEF and functional outcome after IS (P>0.05). Sensitivity analyses yielded no evidence of directional pleiotropy or substantial heterogeneity. Multivariable MR analyses demonstrated that, after adjustment for insomnia, smoking initiation, waist-to-hip ratio(WHR) or depression, both COPD and PEF exhibited a causal association with adverse functional outcome following IS.

**Conclusion:** Our research indicated that at the level of genetic causality, no significant associations were observed between lung function indices (FEV1, FVC, FEV1/FVC, PEF) or COPD and functional outcome following IS. Our discovery may challenge the conventional notion that “improving lung function itself can efficiently promote the prognosis of IS function”. Future research needs to further explore other possible causal pathways and consider various factors comprehensively to develop more effective rehabilitation strategies after IS.

## Introduction

Stroke are major causes of morbidity and mortality worldwide.^1^ Approximately 12 million new stroke cases occur globally each year, with 6.55 million deaths attributed to stroke. The comprehensive economic loss caused by ischemic stroke (IS) is estimated at approximately $891 billion per year.^2,3^ In this context, the incidence rate, disability rate, mortality rate and recurrence rate of stroke in our country are all at high levels, which imposes a heavy economic and social burden on society and families. Furthermore, the prevalence and mortality rates are showing an increasing trend year by year.^4^ The proportions of stroke survivors left with varying degrees of motor dysfunction, cognitive impairment and aphasia are 80%, 78.7% and 30%, respectively. Approximately 50% of patients are unable to independently perform daily functional activities three months after the onset of a stroke and up to 40% of patients will face the harsh reality of moderate to severe disability.^5–8^ IS significantly affects the quality of life of patients, hinders their reintegration into society and imposes unbearable emotional and financial burdens on their families.^9,10^ It is therefore crucial to investigate the factors associated with the improvement of functional outcome after IS.

Lung function tests are primarily used to assess the patency of the airways and the size of lung capacity.^11,12^ They hold significant clinical value for the early detection of lung and airway lesions, as well as for evaluating the severity of diseases and their functional outcome.^13–15^ Among others, the main indicators of lung function include: forced vital capacity (FVC), forced expiratory volume in one second (FEV1), the ratio of FEV1 to FVC (FEV1/FVC), the ratio of FEV1 to maximum vital capacity (FEV1/VC MAX) and peak expiratory flow (PEF). The functional outcome of stroke are influenced by various factors, including the type of stroke, age at onset, severity of the condition, rehabilitation training, the psychological state of the patient, lifestyle habits and social support.^16,17^ Research had found a close connection between lung function and stroke, with common genetic overlap between IS and lung function.^18^ Furthermore, individuals with impaired lung function had an increased incidence of IS.^19^ The study of Choi suggested that a 4-week program of comprehensive diaphragm training for patients with acute stroke results in significantly greater improvements in lung function and respiratory muscle strength compared to the control group, indicating that diaphragm training may reduce respiratory complications following a stroke.^20^ The elevated levels of inflammatory factors in the bodies of chronic obstructive pulmonary disease(COPD) patients not only affect lung function but also impair the recovery of brain tissue through damage to the blood-brain barrier, thereby delaying functional outcome.^21^ Given those rational, it is well accepted that improvement of lung function can exert beneficial effects on the functional outcome after IS. However, a few investigations reported a different finding. For instance, Britto observed that following inspiratory muscle training (IMT) in stroke survivors, no statistically significant changes emerged in measures of functional performance or health-related quality of life.^22^ Likewise, Vaz demonstrated that although six weeks of IMT improved respiratory muscle strength in IS patients, no corresponding gains were identified in maximal inspiratory pressure, 6-minute-walk-test distance, activities of daily living or quality of life scores.^23^ Therefore, can the enhancement of lung function truly be beneficial to the functional outcome after ischemic stroke? Further rigorous investigation may still be required.

The Mendelian randomization (MR) method utilizes genetic variations and other non-experimental data as instrumental variables (IVs).^24^ MR avoids confounding and reverse causation issues present in traditional observational studies, enhancing the availability of genetic association resources.^25–27^ The method of MR has been employed to investigate aspects related to functional outcome after stroke, as well as the effects of lung function and COPD on other diseases.^28–31^ However, there is a gap in MR studies regarding the associations between lung function, COPD and functional outcome after IS. Therefore, this study aims to utilize univariate MR methods to explore the genetic associations between lung function, COPD and functional outcome after IS, providing evidence at the genetic level.

## Materials and Methods

### Study Design

In this study, we used lung function indicators and COPD as IVs, where the lung function indicators include FEV1, FEV1/FVC, FVC and PEF. The functional outcome of IS would be considered as the outcome variable. A two-sample Mendelian randomization approach was employed to examine the causal association. The research design is shown in Figure. 1.

**Figure 1.**
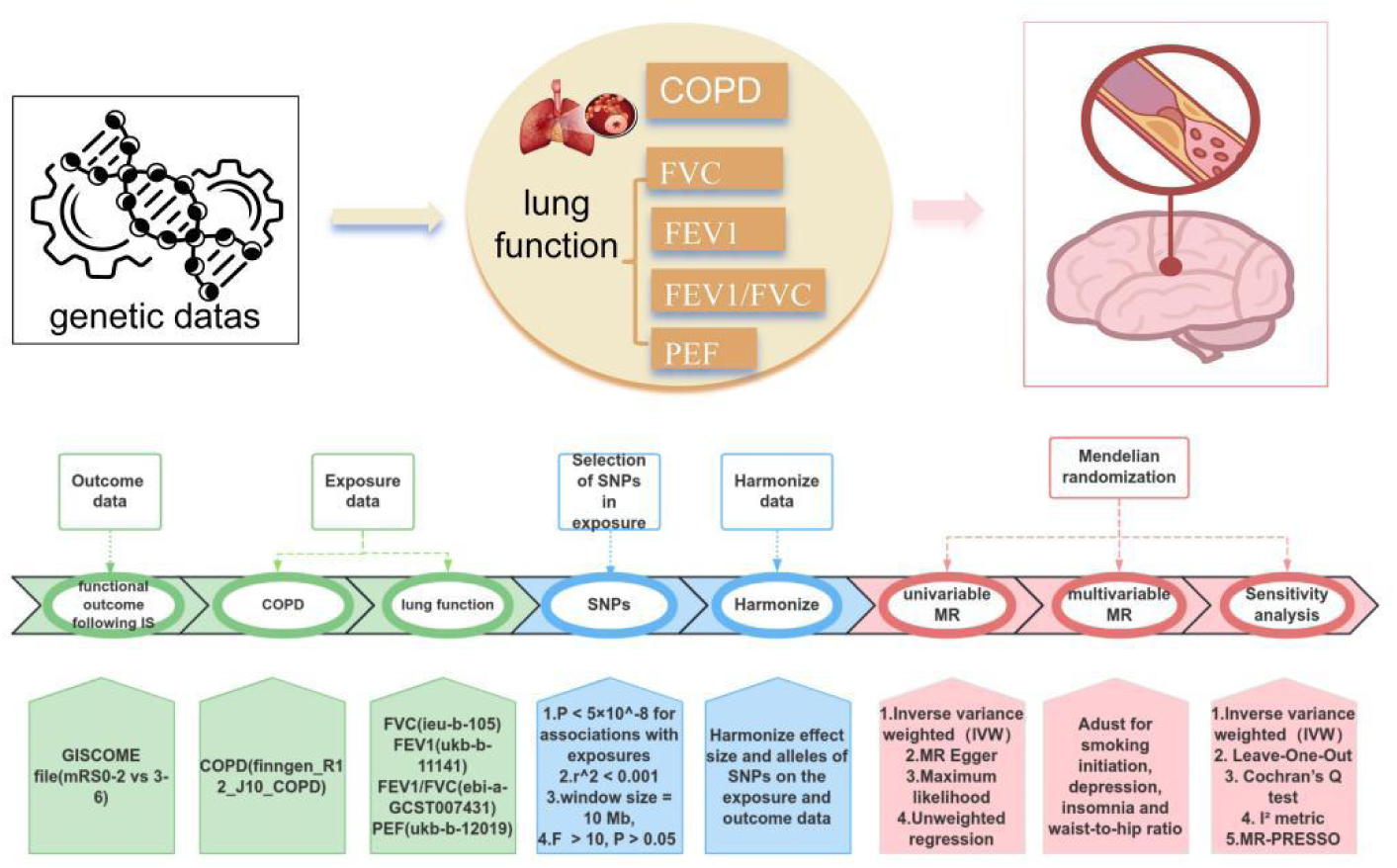
The study design, data sources, and flowchart in the present MR study.

### Outcome data sources

We obtained Genome-wide Association Study (GWAS) result files (mRS 0-2 vs 3-6) related to functional outcome after IS from the Genetic of Ischemic Stroke Functional Outcomes (GISCOME) network (https://kp4cd.org/node/391). This network encompasses 12 studies from Europe, Australia and the United States, involving a total of 6021 patients of European ancestry.^32^ In these studies, the modified Rankin Scale (mRS) score was assessed both as an ordinal variable and as a binary variable, specifically including two comparison forms: mRS 0-2 vs 3-6 and mRS 0-1 vs 2-6. In the GISCOME study,^33^ a total of 3741 patients achieved good functional outcome (mRS 0-2) after IS, whereas 2280 patients experienced poor functional outcome (mRS 3-6). For this analysis, we employed both forms of the 3-month mRS: ordinal mRS and dichotomous mRS (mRS 0-2 vs 3-6).

### Exposure Data

To investigate the genetic associations of lung function, COPD and IS, we obtained relevant data of lung function and COPD from two data resources. Firstly, we accessed the lung function-related GWAS results files (FVC, ieu-b-105; FEV1, ukb-b-11141; FEV1/FVC, ebi-a-GCST007431; PEF, ukb-b-12019) from the IEU OpenGWAS project in the openGWAS summary database (https://gwas.mrcieu.ac.uk/). The GWAS analyses were adjusted for factors such as age, age squared (age^2^), height and smoking status. Additionally, we retrieved the genetic association summary data (finngen_R12_J10_COPD) for COPD from the latest data release of the national genetic factors data repository of the FinnGen project in Finland (https://www.finngen.fi/en). This dataset comprises 433208 samples, with 24138 cases and 409070 controls.

### Selection of IVs

The selection of IVs was completed through a series of quality control procedures: First, among the exposure factors, single-nucleotide polymorphisms(SNPs) with a significance level of *P* < 5 × 10^−8^ were selected. Second, to ensure the independence of each IV, SNPs in linkage disequilibrium were removed based on a r^2^ > 0.001 criterion within a window size of 10,000 kb. Third, SNPs with an *F* statistic greater than 10 were retained to mitigate potential bias caused by weak IVs. The calculation formula for the *F* statistic is: *F* = (R^2^ × (n – k – 1)) / [k × (1 – R^2^)], where R^2^ represents the proportion of variance explained by the phenotype, n denotes the effective sample size and k indicates the number of genetic variations. Final, SNPs that exhibited significant associations with the outcomes of interest (*P* < 0.05) were excluded to prevent pleiotropic effects.

### Statistical analysis

All statistical analyses were performed using R version 4.2.2 (R Foundation for Statistical Computing, Vienna, Austria). Before the analyses of the causal effects, we firstly performed harmonization of the IVs and outcome variables using the ‘TwoSampleMR’ package. This process involved correcting effect directions and standardizing data formats, thereby ensuring the consistency of genetic association data and laying the foundation for subsequent analyses. The inverse variance weighted (IVW) method served as the primary statistical approach, calculating the overall estimate of genetic effects through a meta-analysis of the Wald estimates from each SNP.^34^ Specifically, we weighted the effect sizes of all SNPs by the inverse of the standard error of each SNP (1/SE²) to obtain an overall effect estimate. Furthermore, to assess the robustness of the study results, the weighted median, Two-Sample Maximum Likelihood and MR-Egger methods^35–37^ were used as auxiliary references for the evaluation of the association between the lung function and the functional outcome following IS. Causal effects were expressed as odds ratios with 95% confidence intervals (CIs). The statistically significant association is defined to be *P* < 0.05.

The MR-Egger regression method was used in the multi-IV pooled-level MR test to evaluate whether there was pleiotropy in the IVs. To further determine whether the intercept of the MR-Egger regression is indeed significant or a false positive, the Mendelian randomization pleiotropy residual sum and outlier (MR-PRESSO) was introduced because MR-PRESSO is shown to be more accurate than the MR-Egger regression. Meanwhile, heterogeneity across instruments was evaluated with the I ² metric and Cochran’ s Q test, and a leave-one-out sensitivity check was carried out to determine whether any single SNP disproportionately influenced the pooled estimate.

Moreover, we also conducted multivariable MR analyses to further explore the associations between lung function, COPD and functional outcome following IS by adjusting for the effects of potential confounders. The potential confounders include smoking initiation, depression, insomnia, and waist-to-hip ratio (WHR), which have been shown to be highly associated with both the lung function and the functional outcome after IS.

## Results

### Genetic IVs

According to the selection criteria for IVs, a total of 856 SNPs were identified as IVs for four lung function indicators and COPD in the analysis using the ordered mRS score and binary mRS score. Detailed information regarding the selected IVs can be found in Tab. 1.

**Table 2.** The univariable MR for assessing the causal effect of COPD and lung function on functional outcome of IS.

| Exposures | Method | No. of SNPs | OR (95% CI) | P value |
| --- | --- | --- | --- | --- |
| COPD | Inverse variance weighted | 20 | 1.15 (0.88,1.51) | 0.316 |
|  | MR Egger | 20 | 1.85 (1.03,3.32) | 0.054 |
|  | Weighted median | 20 | 1.44 (0.97,2.14) | 0.073 |
|  | Maximum likelihood | 20 | 1.15 (0.88,1.52) | 0.303 |
| FVC | Inverse variance weighted | 254 | 0.97 (0.66,1.44) | 0.896 |
|  | MR Egger | 254 | 0.70 (0.24,2.02) | 0.512 |
|  | Weighted median | 254 | 1.15 (0.66,2.01) | 0.619 |
|  | Maximum likelihood | 254 | 0.97 (0.66,1.44) | 0.896 |
| FEV1 | Inverse variance weighted | 196 | 1.15 (0.75,1.77) | 0.523 |
|  | MR Egger | 196 | 0.92 (0.23,3.73) | 0.905 |
|  | Weighted median | 196 | 1.46 (0.76,2.77) | 0.253 |
|  | Maximum likelihood | 196 | 1.15 (0.75,1.78) | 0.524 |
| FEV1/FVC | Inverse variance weighted | 265 | 1.06 (0.83,1.36) | 0.65 |
|  | MR Egger | 265 | 1.17 (0.64,2.14) | 0.605 |
|  | Weighted median | 265 | 1.16 (0.78,1.70) | 0.464 |
|  | Maximum likelihood | 265 | 1.06 (0.83,1.36) | 0.652 |
| PEF | Inverse variance weighted | 121 | 1.33 (0.73,2.44) | 0.35 |
|  | MR Egger | 121 | 0.21 (0.03,1.72) | 0.148 |
|  | Weighted median | 121 | 1.83 (0.73,4.57) | 0.198 |
|  | Maximum likelihood | 121 | 1.34 (0.73,2.46) | 0.348 |

### Univariable MR analyses

In univariate MR analyses, the estimates of IVW, MR-Egger, weighted median method and two-sample Maximum Likelihood analysis all showed that there was no significant genetic causal relationship between COPD, FVC, FEV1, FEV1/FVC, PEF and functional outcome after IS (Tab. 1, Fig. 2. *P* > 0.05).

**Figure 2.**
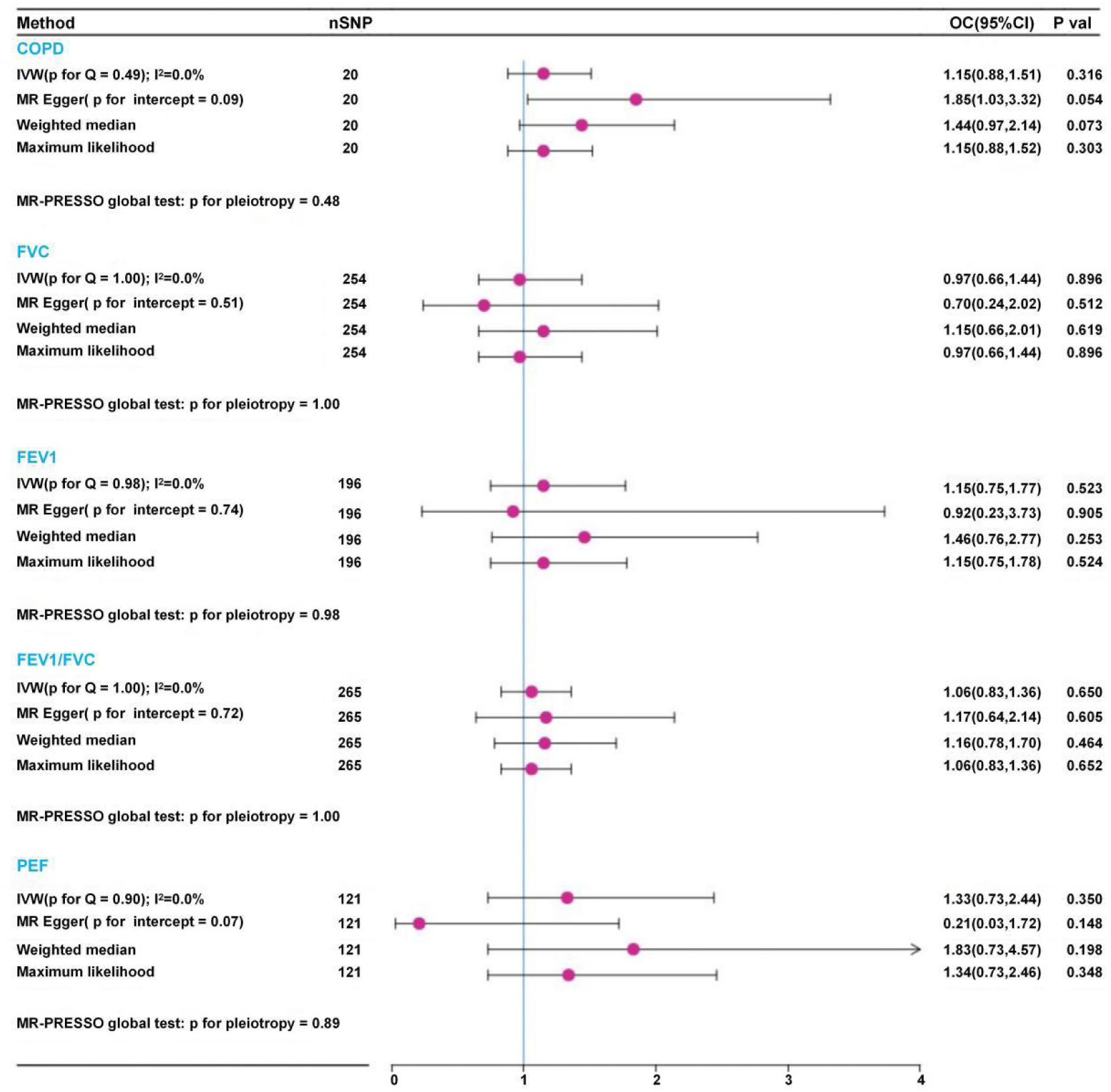
The forestplot of univariable MR for assessing the causal effect of COPD and lung function on functional outcome of IS.

### Sensitivity Analyses

In our sensitivity analyses, heterogeneity was examined using the I² statistic (*P* > 0.05) and Cochran’ s Q test (Q < 15.186, *P* > 0.05), indicating no significant heterogeneity among the instrumental variables (IVs) for lung-function measures (FEV1, FVC, FEV1/FVC, PEF) or for COPD. The MR-Egger regression intercept test for directional pleiotropy showed no evidence of potential pleiotropic bias in the MR estimates linking lung function traits and COPD to IS functional outcome (all *P* > 0.05). Correction with MR-PRESSO likewise revealed no significant horizontal pleiotropy (all *P* > 0.05). Both MR-Egger and MR-PRESSO detected no material pleiotropic distortion in any of the other causal estimates.

Leave-one-out sensitivity analyses demonstrated that no single SNP materially altered the overall causal estimates in any of the univariable MR analyses for the lung-function indices (Fig. 2). Funnel plots from the IVW-based MR analyses exhibited generally symmetrical distributions, indicating the absence of marked small-study bias.

### Multivariable MR analyses

Compared with univariable MR findings, multivariable MR analyses in which COPD was adjusted for insomnia, smoking initiation and depression revealed a potential causal effect on functional outcome after IS (Tab. 2; adjusting for insomnia, β: 0.29, SE: 0.11, OR: 1.34, 95%CI: 1.09-1.65, *P* = 0.006; adjusting for smoking initiation, β: 0.28, SE: 0.09, OR: 1.32, 95%CI: 1.12-1.57, *P* = 0.001; adjusting for depression: β: 0.31, SE: 0.12, OR:1.37, 95%CI: 1.08-1.72, *P* = 0.009). Similarly, PEF adjusted for WHR and insomnia showed an analogous causal association with functional outcome following IS (adjusting for WHR, β: –1.33, SE: 0.42, OR: 0.27, 95%CI: 0.12-0.61, *P* = 0.002; adjusting for insomnia, β: –0.68, SE: 0.33, OR: 0.51, 95%CI: 0.27-0.98, *P* = 0.042). These findings indicated a causal relationship between COPD and functional outcome following IS after controlling the effect of insomnia, smoking initiation and depression. PEF also had a causal relationship with functional outcome following IS after controlling the effect of WHR and insomnia.

**Table 2.** The multivariable MR for assessing the causal effect of COPD and lung function on functional outcome of IS (ordinal mRS) after adjusting WHR, smoking initiation, depression.

| Exposures | Condition | $\beta$ | SE | OR (95% CI) | *P* value |
| --- | --- | --- | --- | --- | --- |
| COPD | WHR | 0.00 | 0.17 | 1.00 (0.72,1.39) | 0.98 |
| COPD | Insomnia | 0.29 | 0.11 | 1.34 (1.09,1.65) | 0.006 |
| COPD | Smoking initiation | 0.28 | 0.09 | 1.32 (1.12,1.57) | 0.001 |
| COPD | Depression | 0.31 | 0.12 | 1.37 (1.08,1.72) | 0.009 |
| FVC | WHR | -0.14 | 0.24 | 0.87 (0.55,1.39) | 0.568 |
| FVC | Insomnia | -0.15 | 0.18 | 0.86 (0.60,1.24) | 0.425 |
| FVC | Smoking initiation | -0.06 | 0.21 | 0.94 (0.63,1.41) | 0.76 |
| FVC | Depression | -0.10 | 0.19 | 0.90 (0.63,1.30) | 0.574 |
| FEV1 | WHR | -0.13 | 0.28 | 0.88 (0.51,1.51) | 0.632 |
| FEV1 | Insomnia | -0.07 | 0.21 | 0.94 (0.62,1.43) | 0.762 |
| FEV1 | Smoking initiation | 0.27 | 0.20 | 1.31 (0.89,1.92) | 0.166 |
| FEV1 | Depression | -0.10 | 0.21 | 0.91 (0.60,1.37) | 0.643 |
| FEV1/FVC | WHR | 0.01 | 0.17 | 1.10 (0.73,1.40) | 0.956 |
| FEV1/FVC | Insomnia | -0.16 | 0.11 | 0.85 (0.68,1.06) | 0.147 |
| FEV1/FVC | Smoking initiation | -0.16 | 0.11 | 0.85 (0.68,1.07) | 0.163 |
| FEV1/FVC | Depression | -0.14 | 0.11 | 0.87 (0.70,1.09) | 0.220 |
| PEF | WHR | -1.33 | 0.42 | 0.27 (0.12,0.61) | 0.002 |
| PEF | Insomnia | -0.68 | 0.33 | 0.51 (0.27,0.98) | 0.042 |
| PEF | Smoking initiation | -0.20 | 0.30 | 0.82 (0.46,1.46) | 0.501 |
| PEF | Depression | -0.47 | 0.34 | 0.63 (0.32,1.23) | 0.173 |

## Discussion

It is well accepted that enhancing lung function exerts beneficial effects on the recovery of a wide range of diseases. In this study, we used the MR approach to explore the causal association between lung function, COPD, and functional outcome after IS. By employing genetic variation as IVs, the results of this study revealed no significant genetic causal relationship between lung function (FEV1, FVC, FEV1/FVC and PEF), COPD, and the functional outcome after IS. This study provided evidence at the genetic level that may challenge the commonly held view that enhancing lung function exert beneficial effects on the functional outcome of IS. This finding is significant for re-evaluating the key points in rehabilitation training and the clinical management strategy for patients following IS. It may provide a reference for developing a forward-looking prevention and management strategy that aims to improve the functional outcome following IS.

### The association among lung function, COPD and functional outcome following IS

A substantial amount of observational evidence has consistently indicated that favorable lung function and a low burden of COPD were not only associated with a reduced risk of incident stroke^18,19,38–41^ but may also exert a beneficial effect on functional outcome following IS.^19–21,42–45^ Proposed mechanisms include improved oxygenation, attenuated systemic inflammation, decreased oxidative stress, preserved endothelial function and maintained coagulation-fibrinolysis homeostasis.^21,43,46–48^ Collectively, these studies had reinforced the view that optimizing lung function and controlling COPD represent potentially modifiable therapeutic targets for enhancing functional outcome in patients with IS. However, using MR with genetic variants as IVs to infer causality, our study obtained results that diverge markedly from the above conventional understanding. Univariable MR analyses, including IVW, MR-Egger, weighted median and two-sample maximum-likelihood estimators, uniformly demonstrated no significant causal associations at the genetic level between lung function indices (FEV1, FVC, FEV1/FVC, PEF) or COPD liability and functional outcome following IS assessed by the mRS (all *P* > 0.05). Consequently, genetically predicted baseline lung function or COPD does not appear to materially influence the extent of neurological recovery following IS.

Several potential explanations may account for the negative findings of our univariable MR analyses. First, residual confounding inherent in prior observational studies was likely to have generated spurious associations. Patients with diminished lung function or COPD frequently exhibit a cluster of co-occurring risk factors, such as advanced age, smoking initiation, hypertension, diabetes, obesity, depression, insomnia and others factions.^17^ These factors are themselves strong predictive indicators of poor functional outcome following IS. Consequently, the apparent relationship between lung impairment and adverse prognosis may reflect the aggregate influence of these pervasive confounders rather than an independent causal effect of lung function. Our multivariable MR results lend partial support to this interpretation: after adjustment for WHR, smoking initiation, depression and insomnia, COPD and PEF demonstrated associations with functional outcome following IS. This result suggested that the adjusted variables may constitute more proximal determinants or critical mediators of prognosis, while lung indicators (particularly COPD and PEF) largely serve as proxies for the presence of these confounding factors or their downstream effects. Second, clinical respiratory rehabilitation, such as IMT, may improve respiratory muscle strength.^49,50^ But primarily enhances are dyspnea and exercise tolerance,^51^ without necessarily synchronously augmenting cerebral hemodynamics or neural plasticity.^52,53^ The recovery of neurological function is more dependent on the salvage of the ischemic penumbra,^54^ reperfusion time and neuronal plasticity, rather than the lung function itself. Therefore, improvements in lung function and COPD do not have a significant impact on the mRS. Third, MR analysis relies on genetic variations that are closely associated with exposure factors.^55^ Although this study selected genetic variations that are strongly linked to lung function indicators and COPD as IVs, these genetic variations may not fully represent the complexity of lung function and COPD. Furthermore, COPD has an intrinsic relationship with lung function measures such as FVC, which may impact the estimation of causal effects. Fourth, The impact of lung function and COPD on functional outcome following IS may involve various biological mechanisms, including inflammatory responses,^56^ oxidative stress^40,57^ and endothelial function.^58^ MR analysis primarily focuses on the influence of genetic variations on exposure factors and these genetic variations may not directly participate in the aforementioned biological mechanisms, leading to insignificant causal effects.

### Strengths and Limitations

Through the comprehensive analysis process described above, we have not only validated the causal relationship between lung function, COPD and functional outcome after IS from multiple perspectives, but also enhanced the credibility and robustness of our research findings through cross-validation using various statistical methods. MR provides evidence at the genetic level to make adjustments to the perception of authority.

However, this study also had some limitations. First, MR analysis relies on genetic variation as tools for gene prediction, which may not fully represent the complexity of lung function and COPD. Second, the data used in this study was regionally biased, primarily based on data from European ancestry populations. The applicability to other racial groups may be limited. More extensive database support is needed to address the requirements of different regions and ethnicities. Third, this study analyzed the association between the core indicators of lung function (FEV1, FVC, FEV1/FVC, PEF) and the functional outcome following IS. Moreover, there are many other indicators of lung function that were not included in this analysis due to the limitations of current GWAS data. Therefore, in clinical practice, while monitoring and managing the lung function and COPD status of IS patients is crucial for their overall health and rehabilitation participation, the effectiveness of interventions targeting lung function and COPD as a core causal strategy for directly improving stroke functional prognosis may be limited. It is important to consider the concurrent management of confounding factors such as smoking, WHR, depression and insomnia. Our study was merely based on genetic level predictions and future multicenter, high-quality RCTs are needed for validation.

## Conclusion

The results of this study indicated that at the level of genetic causality, no significant associations were observed between lung function indices (FEV1, FVC, FEV1/FVC, PEF) or COPD and functional outcome following IS. Our discovery may challenge the conventional notion that “improving lung function itself can efficiently promote the prognosis of IS function”. This finding offered genetic-level evidence for reevaluating the role of lung function in the functional outcome following IS. It may serve as a reference value for optimizing stroke rehabilitation strategies in clinical practice. However, considering the limitations in the design of our study, the results of this MR study should be interpreted with caution.

## Data Availability

All data in this study were obtained from open access platforms. The IS outcome data is sourced from the GISCOME file (mRS 0-2 vs 3-6) available at https://kp4cd.org/node/391; the GWAS results related to lung function (FVC, ieu-b-105; FEV1, ukb-b-11141; FEV1/FVC, ebi-a-GCST007431; PEF, ukb-b-12019) are sourced from the IEU OpenGWAS project (https://gwas.mrcieu.ac.uk/). The COPD data (finngen_R12_J10_COPD) is sourced from https://www.finngen.fi/en. The full set of SNP-exposure and SNP-outcome associations used in the Mendelian randomization analysis is provided in Supplementary materials.

## Acknowledgements

We thank the FinnGen, OpenGWAS and GISCOME network for providing summary data for the analyses.

## Authors’ contributions

All authors contributed to analysis, interpretation and final manuscript preparation. WTS and JW contributed to study concept, rationale and initial manuscript drafts. ZYY, FP, ZDG, YY, JL and LYZ contributed to data acquisition and data analysis. LYZ and GXX contributed to project management and personnel arrangement. All authors read and approved the final manuscript.

## Availability of data and materials

All data in this study were obtained from open access platforms. The IS outcome data is sourced from the GISCOME file (mRS 0-2 vs 3-6) available at https://kp4cd.org/node/391; the GWAS results related to lung function (FVC, ieu-b-105; FEV1, ukb-b-11141; FEV1/FVC, ebi-a-GCST007431; PEF, ukb-b-12019) are sourced from the IEU OpenGWAS project (https://gwas.mrcieu.ac.uk/). The COPD data (finngen_R12_J10_COPD) is sourced from https://www.finngen.fi/en.

## Ethics approval and consent to participate

Ethical approval and consent was not specifically for this study as we used summary data that is publicly available.

## Conflicts of interest

The authors have no financial conflicts of interest.

## Abbreviation

COPD: chronic obstructive pulmonary disease
IS: ischemic stroke
FVC: forced vital capacity
FEV1: forced expiratory volume in one second
FEV1/FVC: the ratio of FEV1 to FVC
FEV1/VC MAX: the ratio of FEV1 to maximum vital capacity
PEF: peak expiratory flow
MR: Mendelian randomization
IVs: instrumental variables
mRS: modified Rankin Scale
GISCOME: Genetic of Ischemic Stroke Functional Outcomes
GWAS: Genome-wide Association Study
SNPs: single nucleotide polymorphisms
IVW: inverse variance weighted
MR – PRESSO: Mendelian randomization pleiotropy residual sum and outlier
WHR: waist-to-hip ratio
IMT: inspiratory muscle training
CIs: confidence intervals

## Notes

### Competing Interest Statement

The authors have declared no competing interest.

### Funding Statement

No funding

